# A standalone approach to utilize telomere length measurement as a surveillance tool in oral leukoplakia

**DOI:** 10.1101/2020.09.25.20193946

**Authors:** Jagannath Pal, Yogita Rajput, Shruti Shrivastava, Renuka Gahine, Varsha Mungutwar, Malti Sahu, Tripti Barardiya, Ankur Chandrakar, Pinaka Pani R., Shovana S. Mishra, Hansa Banjara, Vivek Choudhary, Pradeep K. Patra, Masood A. Shammas

## Abstract

Oral Squamous Cell Carcinoma (OSCC) is often preceded by white patch, called oral leukoplakia (OL). Assessing relative telomere length (TL) in OL could be a predicting biomarker. Due to high variability and lack of universal reference, there has been a limited translational application of TL. Here, we describe an approach of evaluating TL using paired PBMC as internal reference and demonstrate its translational relevance. Oral brush biopsy and paired venous blood were collected from 50 male OL patients and 44 male healthy controls. Relative TL was measured by qPCR. TL of each OL sample was normalized to paired PBMC sample (TL ratio). Mean TL ratio in healthy controls with high risk oral habits was shorter than those who did not have these habits (1.093±0.411 and 1.253±0.296, respectively; p=0.071). In OL patients, the mean TL ratio was not only significantly shorter in the patch but also in distal normal oral tissue (0.971±0.317, p=0.0002 and 0.896±0.284, p=0.00001, respectively), relative to healthy control without high risk oral habit. Based on the TL ratio, we proposed a classification of OL into four subgroups. Dysplastic pathology was frequently associated with a subgroup having normal TL ratio at the patch while significantly shorter TL ratio at paired normal distal site. The approach of analyzing TL attrition of oral mucosa, eliminating requirement of external reference DNA, will enable the TL data universally comparable and provide a useful marker to define high risk OL group for follow-up program. Larger studies will further validate the approach and its broader application in other pre-malignant conditions.

## Introduction

Oral mucosa is the first gateway between internal and external environment^1^. As a result, it is constantly exposed to a variety of toxic substances. High risk oral habits such as chewing tobacco products, smoking and drinking alcohol can be the source of high concentrations of toxic substances for oral mucosa. These agents, either separately or in combination with other substances/factors, can also increase oxidative stress within the oral mucosa. Oxidative stress is associated with increased DNA damage which can contribute to genomic rearrangements and instability, posing a risk for malignant transformation of oral mucosa. This risk could be increased in the individuals who develop unhealthy habits such as chewing tobacco products, smoking and drinking alcohol^2,3^. Chronic exposure of the high risk factors leads to oral potential malignant disorder (OPMD) called oral leukoplakia, a white patch-like oral lesion, diagnosed by exclusion of other known oral diseases that do not carry increased risk for cancer^4,5^. As this condition may progress to cancer, there is a need to identify molecular changes which may predict progression. So far, there is no universal molecular marker which can be utilized in clinical practice to predict the progression of disease. In India, the high incidence of oral cancer could be attributed to oral high risk habits which are wide-spread in the population and contribute to precancerous lesions^6-8^.

Each chromosome end is protected by a cap-like DNA protein structure called telomere^9^. Human telomeres consist of conserved telomere sequence repeats (TTAGGGn) with a single-strand 3′G-rich overhang. A set of proteins bind to these sequences and form a cap-like structure, protecting chromosomal ends from degradation and fusion with each other. Telomere length is maintained by an enzyme “telomerase” which is expressed in germ cells but tightly suppressed in most somatic cells^10,11^. In somatic cells, telomeres progressively shorten with each cell division till it reaches a critical limit below which the cells undergo replicative senescence^12,13^. TL can determine the lifespan of an organism. The rate of telomere shortening, which determines the pace of aging, is impacted by a variety of factors including genetics, environment, lifestyle and socioeconomic status. Premature or accelerated TL shortening is associated with increased risk of many age-related diseases as well as cancer^14-16^.

One of the pre-requisites for perpetual proliferation of cancer cells is the acquisition of ability to maintain telomeres. At least in 85% of cancers the telomere length is maintained by telomerase whereas in rest of the cancers, it is maintained by a recombination-mediated pathway called alternative lengthening of telomeres (ALT)^17,18^. If mitogenic signalling is activated in response to environmental insult/s, it can make the somatic cells proliferate continuously leading to telomere shortening below a critical limit. This leads to a crisis-like state in which activation of DNA double strand break (DSB) response may result in chromosomal end fusion and genomic instability^19-21^. Certain environmental factors (such as radiation) may also directly damage telomeres and accelerate the rate of TL shortening. Following crisis, while most of the cells undergo replicative senescence or death, a fraction of cells are rescued from the crisis, either by expressing telomerase or by activating ALT pathway^22^, leading to oncogenic transformation. Therefore we propose that TL can potentially be used for surveillance of individuals with high risk habits or in pre-cancerous conditions like oral leukoplakia. Since excessive telomere shortening is associated with genetic instability, it has strong potential to be used as a molecular marker to predict progression of precancerous conditions.

A wide range of methods have been developed to measure TL. These include terminal restriction fragment (TRF) analysis using hybridization of digested DNA with telomere sequence probes, fluorescent in situ hybridization using peptide nucleic acid (PNA) probes, single telomere amplification and blotting (STELA) and evaluation of relative TL by qPCR described by Cawthon et al., 2002. All these methods have unique advantages as well as limitations^23-25^. A distinct feature of qPCR method is that only small amount of DNA is required and the assay can be performed in high-throughput format and suited for large epidemiological studies. This type of assay is also ideal for clinical utilization. Recently, the qPCR-based absolute TL measurement has also been described by O’Callaghan et al.^24^. However, in spite of years of research and continuous improvement of technology of measuring TL, the implementation of TL measurement in regular clinical practice remains far from reality. This is because a number of factors such as age, gender, genetics, stress and lifestyle can impact TL^14^. In addition, the method/s used for sample collection, storage, DNA preparation and TL determination can also introduce variability in results^6^. Moreover, relativeTL is usually expressed as a ratio of TL of an appropriate reference DNA. As the reference DNA also varies from study to study or laboratory to laboratory, it is difficult to compare the TL from one study with other. Moreover, there is also batch effect on TL during DNA preparation. A recent study on reproducibility of TL measurement in different laboratories across the globe suggests high inter-laboratory coefficients of variation (CVs) which were about 10% and 20% for Southern blotting and qPCR, respectively^27^. There is also large variation of TL due to ethnicity, making it difficult to analyze TL data in mixed multicultural population studies^28^. In some studies, the TL in pathological tissue was compared with that in adjacent healthy tissue^31-35^ to eliminate the impact of other variables or batch effect. However, this may not be ideal because adjacent tissue could also be abnormal if etiological exposure affected a large area or whole organ^34,35^.

In this regard, a distant normal tissue may be a better internal control to eliminate impact of variables and batch effect. Every tissue undergoes variable number of cell divisions and there is also variation of TL in different tissues within an individual. However, physiological shortening occurs at equivalent rate and there is a correlation of TL between different tissues from the same individuals^36-38^. Recently, Finnicum et al. (2017) also showed that TL of PBMC and buccal mucosa are significantly correlated^39^, suggesting the similarities in telomere dynamics between two tissue types within an individual. So, it would be quite feasible to express TL of a tissue of interest relative to TL of an easily accessible distant tissue, preferably PBMC. This should eliminate/minimize the impact of all variables mentioned above and allow utilization of TL as a clinical marker. Additionally, the use of paired PBMC sample will also eliminate the need of an external reference DNA which usually varies from laboratory to laboratory and study to study.

Though many studies have been carried out on TL dynamics in oral cancer^31,32^, information about TL in precancerous stages of oral mucosa is lacking. Recently, using Q-FISH, Aida et al. reported shorter telomeres in high risk ortho-keratotic dysplasia (OKD)-type leukoplakia and in adjacent healthy tissue relative to healthy control^40^. However, the authors in this study did not address the limitation discussed above. Moreover, so far there is no report on TL status in OL at distal apparently healthy oral mucosa.

In this study, we explored the feasibility of utilizing TL of oral mucosa normalized to TL of PBMC of the same subject as a clinical marker for OL. Using this approach, we determined the normal range of TL in healthy subjects and show that telomeres are not only shorter in OL patches but also in oral mucosa of healthy volunteers with high risk oral habits. Telomere shortening was also observed in apparently distal healthy mucosa (anatomical opposite site) of OL patients, indicating that adjacent tissue may not be an appropriate internal control. Therefore, TL in oral mucosa relative to healthy distant tissue such as PBMC should be used to assess pathology and disease outcome in oral potentially malignant disorders. Finally, we propose a classification of OL patients based on combination of normal or short TL in patches as well as in paired distal normal oral mucosa and show its clinicopathological relevance.

## Results

### Demographic profile of healthy controls and leukoplakia patients

Total 66 OL patients and 73 healthy controls (HC) were enrolled for the study. Clinical and demographic parameters of the 60 OL patients have been published earlier^41^. As number of female OL patents were very low and females also behave differently to the exposure of high-risk oral habits, in this pilot study, we only analyzed relative telomere length in male OL patients and compared that with the corresponding male healthy control group. Due to insufficient sample quantity or low quality of DNA, 50 OL and 44 healthy control samples were processed for TL analysis. Mean ages of OL patients and healthy controls were 42.24±12.08 years (range 20-67 years) and 36.36±13.47 years (range 20-69 years) (Table 1), respectively. Out of 44 healthy donors, 25 had no recognizable high-risk oral habit whereas 19 had one or more high risk oral habits which include chewing tobacco, smoking tobacco, and alcohol consumption. Oral habit was considered to be of high risk if it was carried out regularly at least for 2 years consecutively. In case of multiple oral habits, the duration of the longest regular habit was considered. Mean duration of the habit in the high-risk healthy control group was 10.13 ± 8.34 years. Among the OL patients, 47 had high risk oral habit/s and 3 had no high risk oral habit. Mean duration of the habit in the high risk OL group was 15.47±11.40 years. Detail of age distribution of the subgroups of HC and OL patients and duration of high risk habits are shown in Table 1 and Supplementary Table 3, respectively.

**Table 1.**
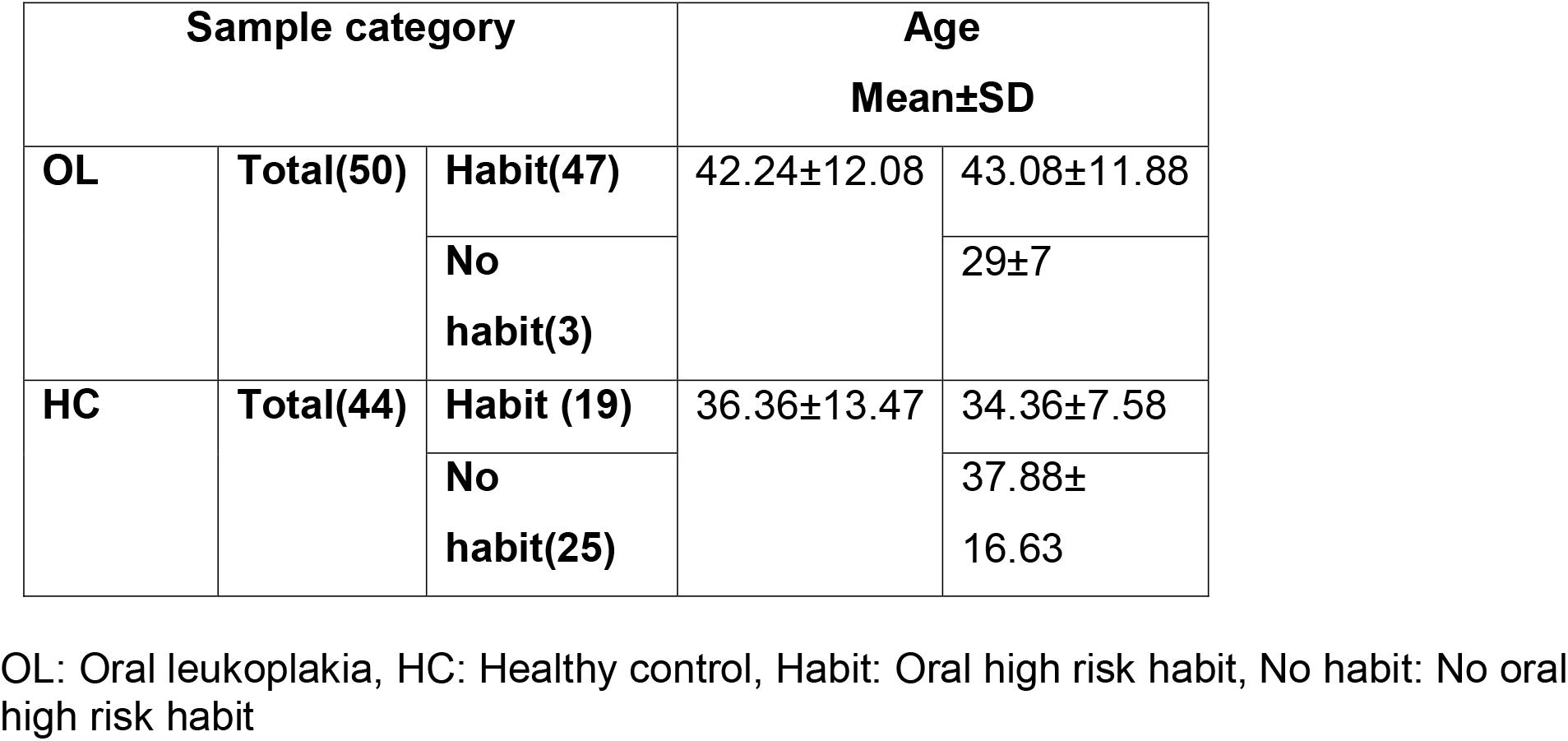
Age distribution among OL patients and healthy controls.

| Sample category |  |  | Age<br>Mean±SD |  |
| --- | --- | --- | --- | --- |
| OL | Total(50) | Habit(47) | 42.24±12.08 | 43.08±11.88 |
|  |  | No habit(3) |  | 29±7 |
| HC | Total(44) | Habit (19) | 36.36±13.47 | 34.36±7.58 |
|  |  | No habit(25) |  | 37.88±16.63 |

OL: Oral leukoplakia, HC: Healthy control, Habit: Oral high risk habit, No habit: No oral high risk habit

### Evaluation of relative TL of oral mucosa in reference to paired PBMC sample

To evaluate TL as a marker for surveillance in OL patients, we used paired normal PBMC sample as internal reference and analyzed ratios of TL in paired samples of oral mucosa and normal PBMC from the same patient. This parameter is mentioned as ‘TL ratio’ in this manuscript. First, we determined the ‘TL ratio’ (indicated as O/M) in the paired samples of oral mucosa and normal PBMC from healthy individuals. Frequency distribution of TL ratio of Healthy controls (total with and without high risk habits) has been shown in Figure 1A. Overall, mean TL ratio of all healthy subjects was 1.18±0.35 (95% CI: 1.076-1.292). The mean TL ratios in healthy male subjects without and with high risk habits were 1.25±0.29 (95% CI: 1.130-1.375) and 1.09±0.41 (95% CI: 0.897-1.292), respectively (Table2, Figure 1B). Median values of both the groups were 1.297 and 1.002, respectively. The TL ratio was not significantly correlated with age in either group (Table1), This signifies that the relative TL expressed as O/M ratio is age-neutral. All values of TL ratio in each group were normally distributed. Test of normality of each group is shown in Supplementary Table 1. As we are only evaluating if the high risk group has shorter TL ratio than no risk habit (healthy) control, one-tailed t test was applied for determining significance of mean differences between the groups. However, we did not observe any statistically significant difference of mean between high risk and no high-risk oral habit groups (p=0.071) (Table 1). This could be due to small difference of means at the given sample size.

**Figure 1:**
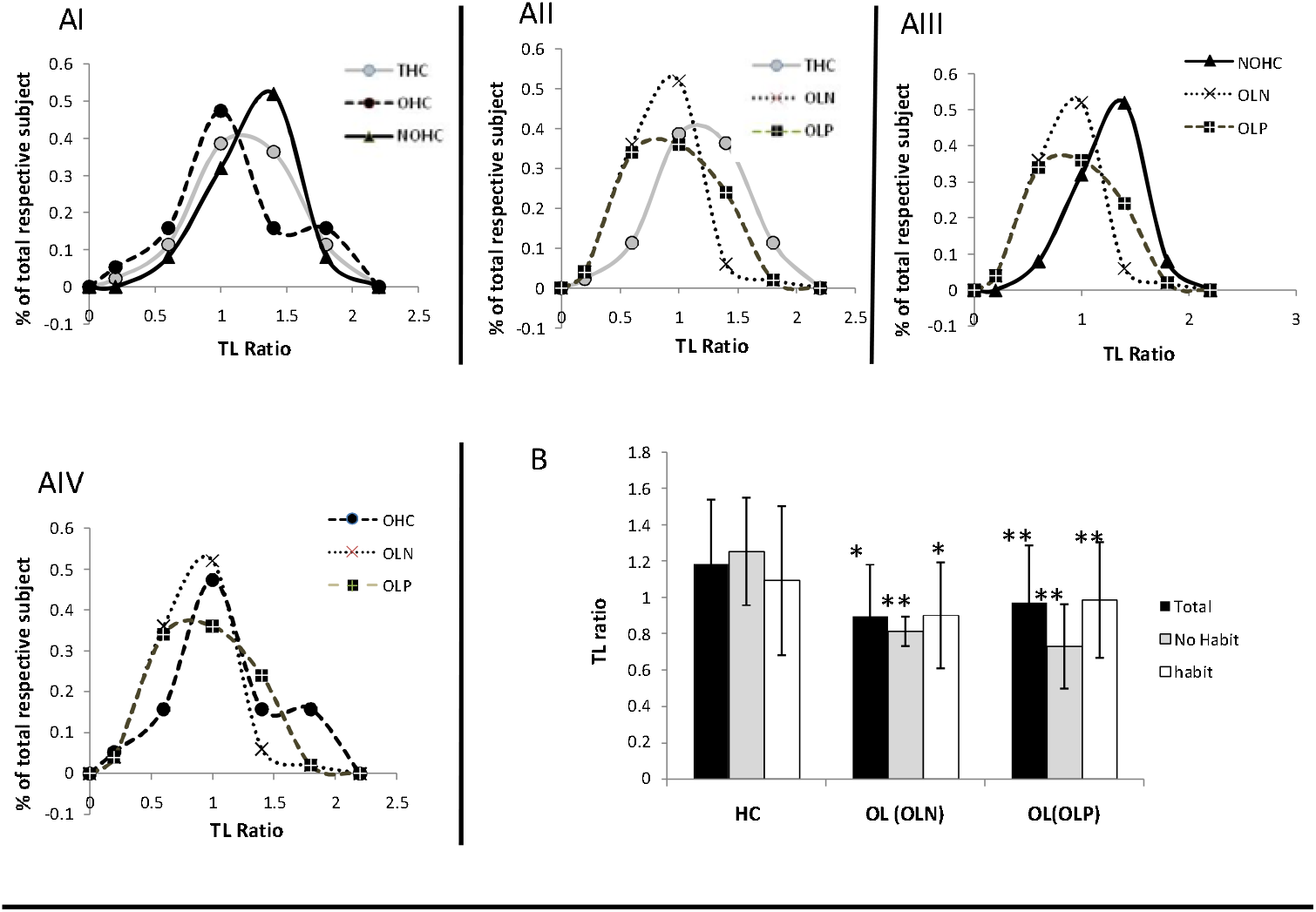
Frequency and mean distribution of TL ratio (O/M ratio) in oral mucosa of the study groups. A. comparison of frequency distribution curves of OL patients with healthy controls. (I) comparative distribution of THC, NOHC and OHC, (II) comparative distribution of OLN, OLP and THC, (III) comparative distribution of OLN, OLP and NOHC, (IV) comparative distribution of OLN, OLP and OHC. HC: healthy control, THC: total healthy control; NOHC: healthy control without oral habit (NOHC); OHC: healthy control with no high risk oral habit; OL: oral leukoplakia; OLP: Oral patch in OL patients; OLN: Paired distal normal mucosa in OL patients. B. Mean distribution of study subjects. Color code indicated in the figure. p<0.5 considered statistical significant. * p Significant when compared to all groups of Healthy controls (total, no habit, habit); ** p significant when compared to total and no habit healthy control,

**Table 2.** Mean TL ratio in healthy control groups and high risk oral habit.

| HC |  | TL ratio Mean ± SD<br>(95% CI) |  | p value | Correlation with<br>Age vs TLR(p) |
| --- | --- | --- | --- | --- | --- |
| Total (44) | OHC<br>(19) | 1.184±0.355<br>(1.076-1.292) | 1.093±0.411<br>(0.897-1.292) | 0.071<br>(OHC<br>vs<br>NOHC) | 0.210(0.388) |
|  | NOHC<br>(25) |  | 1.253±0.296<br>(1.130-1.375) |  | -0.069<br>(0.747) |

HC: Healthy control, NOHC: Healthy control having no oral high risk habit, OHC: Healthy control having oral high risk habit, \*p value is significant at the significance level $p \leq 0.05$ . R: the *r* statistics (measure of Pearson correlation)

Based on these observations, we determined the lower cut off for the TL ratio in healthy male individuals. For males, the TL ratio of 0.765 was set as a threshold (95 percentile of no high-risk habit healthy individuals; one sided Z score 1.645); TL ratio below this could be considered as significantly shorter TL. One-sided percentile was calculated because long TL is not expected to have any pathological significance. Interestingly, 21% (4/19) of healthy males with high risk oral habits had significantly shorter TL in oral mucosa as per our criteria.

### Analysis of TL ratio in oral leukoplakia

TL ratio(O/M) was also assessed in 50 oral leukoplakia samples. Age composition is described in Table 1. Oral leukoplakia predominantly occurs in males with high risk oral habits. Consistently, out of 50 male patients, 47(94%) had recognizable high risk oral habits. For each patient, oral brush biopsy samples were collected from oral leukoplakia patches (OLP) as well from distal visually normal site (OLN), preferably from anatomical opposite site in the oral cavity. All the values of TL ratio in OL patients were normally distributed. Test of normality of each group is shown in Supplementary Table 1. One tailed t test was applied for determining if TL ratio in OL is significantly shorter compared to healthy control groups.

Frequency distribution curves of TL ratio of the OL patient samples compared to corresponding healthy controls have been shown in Figure 1A. Peak TL ratio distributions of both OLN and OLP were clearly in shorter range than corresponding male healthy controls without any high risk oral habit. However, TL ratio distribution of healthy controls with high risk oral habits was very close to distribution of OL samples, (Figure 1A III, IV). Overall, mean TL ratio of OLN and OLP in all OL patients were 0.89±0.28 (95% CI: 0.815-0.976) and 0.97± 0.31 (95% CI: 0.881-1.061), respectively. The same in OL patients with high risk habit were 0.90±0.29 (95% CI: 0.815-0.987) and 0.98±0.31 (95% CI: 0.893-1.080), respectively. In no high risk habit OL patients, the same were 0.81±0.08 (95% CI: 0.608-1.016) and 0.73±0.23 (95% CI: 0.155-1.307), respectively (Table 2, Figure 1B). Though the mean TL ratio in OLP was slightly greater than OLN, it was not significantly different (Table 2). However, a weak statistically significant positive correlation was observed between OLP and OLN (0.389; p=0.005), respectively (Supplementary Table 2). Just like healthy control groups, we did not observe any correlation of TL ratio with age in OL patients (Supplementary Table 2), suggesting the parameter is age neutral.

The mean TL ratios of OLN and OLP in all groups of OL patients (i.e., total high risk and no high risk oral habit groups) were significantly shorter compared to healthy control group having no high risk oral habit (p<0.00001, p=0.00001, p=0.0094 respectively) (Table2). When means of TL ratio in OLN and OLP were compared with the healthy group with high risk oral habits, although both the OLN and OLP (in all the subgroups of OL patients) showed slightly shorter TL ratio, the significance was observed only with OLN but not with OLP (Table2).This might be due to smaller difference of means between two groups and/or small sample size. To verify if duration of high risk exposure may affect TL ratio in OL or in high risk oral habit healthy control groups, we performed Pearson correlation statistics. Interestingly, no statistically significant correlation was observed between TL ratio and duration of oral habits. However, we observed statistically significant moderate correlation between age and duration of oral habit (Supplementary Table 3).

The observation of significant shortening of TL ratio in both high risk and no high risk oral habit OL patients suggests that TL shortening in oral mucosa might be consistently related to pathogenesis of OL irrespective of oral habit. It is yet to be explored if some other etiological factor/s of OL like HPV may contribute in TL shortening of oral mucosa in the absence of any recognizable high risk oral habits. However, we are cautious to draw any conclusion on OL patient group having no high risk oral habit due to very low sample size of this group (Table3).

**Table 3.** Profile of TL ratio in OL patent subgroups.

| **OL Patients (n)** | **Mean TL ratio ±SD (95%CI)** |  | **$\Delta$ Mean, TL ratio ( 95%CI), p** |  |  |  |  |
| --- | --- | --- | --- | --- | --- | --- | --- |
|  | **OLN** | **OLP** | **OLN vs OLP** | **OLN vs NOHC** | **OLP vs NOHC** | **OLN vs OHC** | **OLP vs OHC** |
| **Total (50)** | 0.896±0.284<br>(0.815-0.976) | 0.971±0.317<br>(0.881-1.061) | 0.076<br>(0.044-0.195),<br>0.213 | 0.357<br>(0.217-0.498),<br>**<0.00001\*** | 0.282<br>(0.130-0.433),<br>**0.00020\*** | 0.192<br>(0.025-0.372)<br>0.01299 | 0.123<br>(-0.063-0.308)<br>0.0956 |
| **Habit(47)** | 0.901±0.292<br>(0.815-0.987) | 0.986±0.318<br>(0.893-1.080) | -0.085 (-0.211-0.040);<br>0.178 | 0.352 (0.207-0.497);<br>**0.00001\*** | 0.266<br>(0.113-0.420);<br>**0.00029\*** | 0.193<br>(0.014-0.372);<br>0.0176 | 0.107<br>(-0.081-0.296);<br>0.1291 |
| **No Habit (3)** | 0.812±0.081<br>(0.608-1.016) | 0.731±0.232<br>(0.155-1.307) | -0.081<br>(-0.475-0.314);<br>0.600 | 0.441<br>(0.082-0.799);<br>**0.0090\*** | 0.521<br>(0.155-0.888);<br>**0.0035\*** | 0.282<br>(-0.224-0.788)<br>0.1295 | 0.363<br>(-0.1512 - 0.8769),<br>0.07825 |

Habit: Oral high risk habit, No habit: No oral high risk habit, NOHC: Healthy control having no oral high risk habit, OHC: Healthy control having oral high risk habit, \*p value is significant at the significance level $p \leq 0.05$

### Classification of OL based on TL ratio

Using the cut-off up to 0.76 lower limit of normal TL ratio of oral epithelium, we identified total 22 out of 50 (44%) cases of male OL patients who had significantly shorter relative TL at least in one oral sample (OLP/OLN). Since chronic exposure of oral mucosa to high risk factor/s may contribute to etiology of OL or carcinoma, we investigated TL not only at OL patches but also at distal normal oral mucosa. Based on the established TL ratio threshold values, OL patients categorized into four sub-groups: (A) OLN normal – OLP shorter; (B)OLN shorter – OLP normal; (C)OLN shorter – OLP shorter; (D)OLN normal –OLP normal. The respective percentages of OL patients in the groups A, B, C and D were10% (5/50), 20% (10/50), 14%(7/50) and 56%(28/50), respectively (Table 4)

**Table 4.**
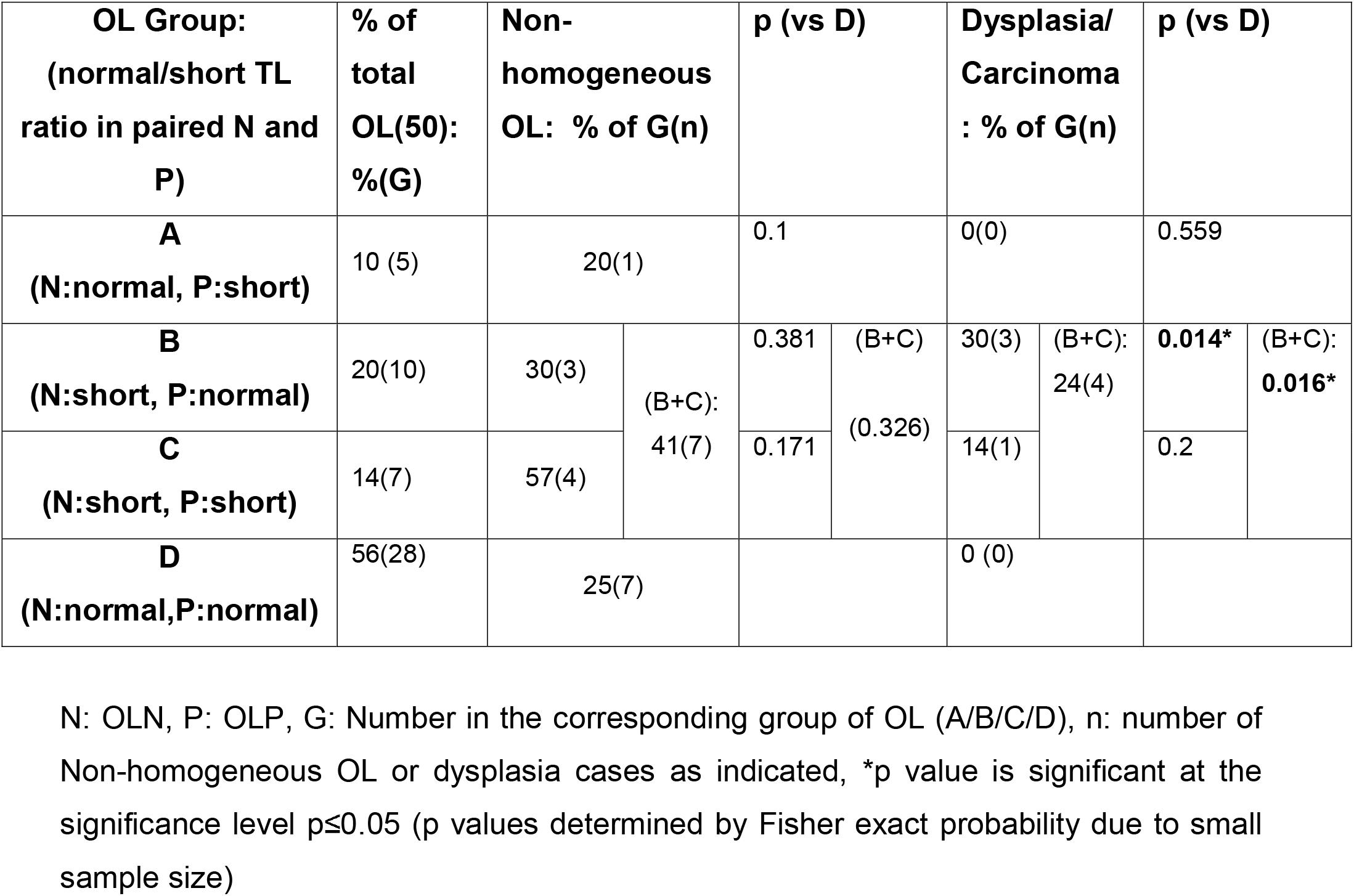
Clinicopathological association with the TL ratio-based subgrouping of OL.

### Clinicopathological association with the TL-based classification of OL

As non-homogeneous OL (NH) is one of the high-risk clinical presentations of OL for malignant transformation^42^, frequency of NH was evaluated in TL-based OL subgroups. NH was defined as a predominantly white or white and red (speckled) and irregularly flat, nodular or verrucous oral lesions^43^. In the present study we identified total 15 NH out of the 50 OL samples. Interestingly, we observed the highest percentage of NH in group C i.e., shorter telomeres in both the OL and adjacent normal samples (57%; p=0.171 compared to group D) followed by B (30%), A (20%) and D (25%). Combining groups B and C, the frequency of NH was 41% (Table 4) suggesting that OL with shorter TL ratio in OLN, irrespective of TL ratio in the paired OLP, may be more frequently associated with NH compared to group D (TL ratio of OLN/OLP within normal range). However,we did not find any statistically significant association of NH with any of the groups relative to group D. Therefore, larger study is needed to determine any statistically significant association.

In this group of samples, only 4 out of 50 male OL patients (8%) were identified to be oral dysplasia or early stage carcinoma (2 dysplasia and 2 carcinoma). We analyzed them as category of bad pathological outcome. When we compared the occurrence of dysplasia/carcinoma in the respective OL groups (A, B, C, D), we observed the occurrence of all dysplasia/carcinoma samples only in groups B followed by C. Group A and Group D both showed 0% occurrence of dysplasia/carcinoma in our study. Combining B and C, the proportion was 24% (Table 4). Corresponding relative risk (RR) and p values for association of dysplasia with the group B and C compared to group D were 18.45 (95% CI: 1.04 - 329.02), p=0.014 and 10.88 (95% CI: 0.488 - 242.24), p=0.20, respectively. Due to small sample size test of association (p) was determined by Fisher exact probability test. The same for Combined B+C was 14.50 (95% CI: 0.829 - 253.697), p=0.016.

As the clinicopathological presentations of non-homogeneous OL, dysplasia, and carcinoma all carry a high risk of transformation into invasive malignancy, we combined all these presentations as a high risk category. With total 18 out of 50 OL cases in this category, the TL ratio at OLN site was significantly shorter in 55.5% cases compared to that in 21.9% cases with low risk OL (homogeneous OL without any dysplasia/carcinoma) (Table 5; p=0.017). This indicates that shorter telomers at OLN are associated with a relative risk 2.43 (95%CI: 1.18-5.00), p=0.0160.

**Table 5.** Distribution of TL ratio in OLN in reference to combined clinicopathological high and low risk groups.

| **TL ratio in OLN** | **NH+D/C**<br>**% of**<br>**total(n)** | **H without**<br>**D/C% of**<br>**total(n)** | **Relative Risk**<br>**RR(95%CI), p** |
| --- | --- | --- | --- |
| **Short** | 55.5(10) | 21.9(7) | 2.43(1.18-5.00),<br>p=0.016* |
| **Normal** | 44.5(8) | 78.1(25) |  |
| Total | 18 | 32 |  |

NH: non-homogeneous OL, D/C: Dysplasia/Carcinoma, n: number of corresponding cases, H: homogeneous OL, \*p value is significant at the significance level $p \leq 0.05$

These observations indicate that the high risk subgroups of OL (NH and dysplasia/carcinoma combined) are more frequently associated with shorter TL in distal normal oral mucosa (OLN), irrespective of TL status of OL patch (group B &C).

## Discussion

In this study, we present an approach to analyze TL in oral mucosa in normal or precancerous conditions, relative to paired PBMC sample from the same patient. This simple approach could eliminate several existing limitations related to utilization of TL as a clinical marker. Using this approach, we were able to segregate OL patients in four groups based on their TL in oral patches and distal normal site and showed its clinicopathological relevance.

The approach was able to detect the pathological changes in TL in oral epithelium even in apparently healthy individuals with high risk habits. Recently, Adalla et al. ^44^ has demonstrated that in patients with inherited bone marrow failure syndromes, the TL in buccal cells correlates with that in blood cells. It indicates that even in a genetic disease involving TL dysfunction, a correlation is maintained within TL of different tissues. Interestingly, overall buccal/blood TL ratio in that study was very close to the overall TL ratio (healthy control) of our study (Table 1). This suggests that utilization of TL of paired PBMC sample as internal control eliminates or minimizes the impact of internal variables such as age, genetic makeup and allows evaluation of the impact of exposure or etiological agent/s being investigated. As the parameter TL ratio is age neutral (no correlation with age), for interpreting data in clinical application, age-adjusted control will not be required thus making the application simpler.

Lower limit for normal male TL ratio (O/M) of oral mucosa was determined 0.76. However, a larger study is required to confirm and refine these numbers. Moreover, it remains to be investigated if TL ratio (O/M) is stable across different populations and regions. If not, the region-specific normal TL values will have to be established.

Although OL patches develop on a focal area, the prior exposure of high risk factors always involves larger areas of oral mucosa. So, to understand the genomic pathology and disease outcome, it is important to evaluate the patch as well as apparently healthy tissue at distant site. So unlike previous studies, we sampled patch area as well as clinically normal mucosa from anatomically opposite side. This approach was able to demonstrate that TL shortening happens not only in patch but also in apparently healthy oral mucosa at anatomically distant site.

Additionally, in our study expressing TL of oral mucosa in OL as proportion to paired PBMC as internal reference provided unbiased unique insight regarding the local molecular pathology of the disease progression. Interestingly, we also observed that overall TL ratio in OL patients was shorter at the normal site (OLN) than the paired patch (OLP). However, more sample size might be needed for observing a statistical strength.

Though we observed shorter TL ratio in no habit group than high risk habit group of OL, we are cautious to conclude this because we were able to register only total 3 OL patients without having any high-risk oral habit. Interestingly, OL in the absence of any high risk oral habit is also more prone to carcinogenic transformation^45^. It may be possible that other etiological factor/s (e.g. HPV infection) affect genome of oral mucosa before appearance of OL in the absence of high risk oral habits.

Paradoxically, we did not observe any correlation of TL ratio with the duration of high risk habit in both healthy control and OL groups. However, as expected the duration of habit strongly correlated with age of the subjects. There might be other internal host factors like genetic susceptibility, nutritional status etc. which interplay with high risk habit to impact telomere attrition.

In our study, only 4 out of 50 OL patients (8%) were identified as dysplasia/carcinoma. This is consistent with a previous study in which screening of a cohort of 2920 OL patients revealed that only 8.6% of cases were dysplasia and 0.72% identified as malignant. Together 9.3% cases were either dysplasia or malignant^49^. However, the occurrence of dysplastic changes and malignant transformation of OL varied grossly. This is because it depends on several factors including the practice of oral high risk habits, duration of exposure in the study population, and genetic background etc. In a meta analysis of 24 studies, a wide range of malignant transformation rate (0.13% to 34.0%) in OL was observed^42^.

Based on combination of normal/short TL in patch and distal normal oral mucosa, we segregated OL patients in 4 subgroups (Table 3). This grouping will help to understand molecular pathogenesis and progression of precancerous conditions. Normal TL in patches in a background of short TL in normal oral mucosa as defined in group B might represent activation or increase in a TL maintenance mechanism at the site of patch. This could be by telomerase or recombination-medicated (ALT) mechanism and could be transient or persistent. Persistent reactivation might be more alarming for risk of developing carcinoma. Higher TL in patches could be indicative of acquiring more survival and proliferating capacity over a background of oral mucosa with severely short TL. Interestingly Sainger et al (2007) has also observed poor prognosis in the patients with higher tumor to adjacent normal tissue telomere length ratio^32^ in oral cancer. A recent report has also shown that shorter relative TL in adjacent normal tissue is associated with severity of tumor stages, CIN and tumor progression in colorectal cancer^30^. There is evidence that normal cells with short telomeres enter senescence stage and secrete several signaling molecules which help neighboring cells to reenter cell cycle by passing the senescence signal, resulting in CIN and tumor progression^46-48^. This suggests that identification of this group with close follow-up prior to clinical tumor formation may reduce incidence of more fatal form of oral malignancy. However, for the purpose of a follow-up program, groups B and C can be combined. This will include 47% (7/15) of non-homogeneous OL and 100% (4/4) of dysplastic/carcinoma in situ OL cases. The fact that all dysplasia samples significantly associated with groups B and C further supports the strength of our approach of identifying high-risk OL patients. Non-homogeneous OL also increases the risk of malignancy^42^. In this study though we observed an increased occurrence of non-homogeneous OL in groups having shorter TL in distal normal site (groups B and C), the association was not statistically significant. However, the classification of homogeneous and non-homogeneous OL is very much subjective and this could be the reason that we were unable to detect any significant association of non-homogeneous OL with short TL groups in distal normal site (B and C). Interestingly, when we combined all the clinicopathological high risk presentations of OL (i.e., non-homogeneous OL and dysplasia/carcinoma combined) in a single category, we observed it is strongly associated with short TL ratio in OLN. This might be due to nullification of any subjective biasness which could occur when clinical and pathological categories are considered as separate high risk group.

Carrying out a follow-up program accommodating a large number of OL cases for early detection of malignant transformation is a huge burden on health service providers. Therefore, the identifyication of potential high risk OL group is essential to reduce the number of patients for follow-up surveillance. Any clinical and histopathological risk assessment also carries a wide intra and inter observer variability^50,51^. The approach of TL evaluation in OL could be used as additional biomarker for evaluating disease progression. Moreover, as punch biopsy is very painful, inconvenient and associated with low patient compliance, a surveillance based on TL ratio will eliminate the need of more frequently invasive punch biopsy as well as will define smaller high risk target group for follow-up program with special attention, which will in turn reduce cost of health care system.

The evaluation of relative TL of oral mucosa normalized to paired PBMC sample is also expected to serve as a standalone and universally comparable clinical marker of oral health and also would be comparable within heterogeneous population without requirement of complicated matched control and reference DNA for every analysis. This approach could also be used for evaluating TL of any solid tissue pathology. It could be applied to other precancerous conditions in esophagus and cervix etc. Additionally, this method could also be useful for any longitudinal study without requirement of simultaneous processing of baseline and follow-up samples for comparison. Other than cancer, the method could also be explored in analyzing sperm telomere attrition in cases of infertility.

## Materials and methods

### Ethical statement

Study was carried out at Multi-Disciplinary Research Unit (MRU), Pt. Jawaharlal Nehru Memorial Medical College, Raipur, Chhattisgarh, India, following approval of the protocol by institutional ethical committee, Pt. J.N.M. Medical College, Raipur, India. All human samples were collected following informed consent from the study subjects.

### Study participants

Patients who attended ENT and dental clinic with oral lesion and clinically diagnosed as leukoplakia and willing to participate in the study were referred to MRU for enrollment. Normal course of clinical management of the patients were not hampered by the study protocol. Patients undertaking any therapy for the lesion, history of other chronic diseases, bleeding tendency, immunocompromised patients and presented with any acute illness were excluded from the study.

### Sample collection

Brush biopsy: Prior to oral brush biopsy, study subjects were asked to rinse mouth twice thoroughly with normal saline. Brush biopsies were performed using small-headed baby toothbrush on the leukoplakia lesion/s (OLP) and on a normal site preferably corresponding opposite side (OLN). In case of healthy control, buccal samples were collected from cheek. Cells were dislodged into PBS, centrifuged, and washed with PBS. Slides were prepared for cytological assessment and rest of the cells were stored at −80°C in 10% DMSO. Blood collection: Blood were collected by venepuncture in EDTA and non EDTA vials.

### Isolation of PBMC

Isolation of PBMC was done using Granulosep GSM and Hisep LSM (HI Media, Mumbai, India) as per manufacturer protocol. Cell pellet was re-suspended in 10% DMSO + RPMI media and stored in aliquots at −80°C till further use.

### DNA isolation and quantization

All paired oral brush biopsy samples and PBMC were processed simultaneously. DNA isolation was done by DNeasy blood and tissue kit (Qiagen, Hilden, Germany) using manufacturer’s protocol with little modification. Proteinase K treatment was carried out over night at 56°C. For elution, column was kept with elution buffer at room temperature for 2 hours before centrifugation. DNA quantization was performed by Quant-iT PicoGreen dsDNA Assay Kit (Invitrogen, Eugene, OR) as per manufacturer’s protocol using Multimode Microplate Reader (TECAN, Mannedorf, Switzerland).

### Relative Telomere length assay by qPCR

Telomere length (TL) measurement was performed using quantitative PCR method as described by O’Callaghan and Fenech with little modifications. Instead of absolute telomere length as described by O’Callaghan et al., we measured relative telomere length (Details in supplementary materials and methods).

### Statistical analysis

Mean, standard deviation, median and Pearson correlation were performed in excel sheet. Kolmogorov-Smirnov Test of Normality, Comparison of mean (t-test), significance of pearson’s correlation, significance of association,(Fisher exact probability due to small sample size) and relative risk were analyzed using the following web based software: MedCalc (htmlhttps://www.medcalc.org/calc/odds_ratio.php) and Social Science Statistics (https://www.socscistatistics.com/tutorials/ttest/default.aspx). Statistical significance was considered if p=< 0.05.

## Data Availability

all data available in the manuscript.

## Acknowledgements

This work was supported by the Department of Health Research, Ministry of Health, India. We also acknowledge Department of Oral Medicine & Radiology, Government Dental College, Raipur for all necessary support. For analytical and bioinformatic support, we would also like to acknowledge Drs. Mehmet K. Samurand Nikhil C. Munshi whose research is supported by Department of Veterans Affairs Merit Review Award I01BX001584-01 (NCM), NIH grants P01155258 and 5P50 CA100707 (NCM, MAS, MKS) and Leukemia and Lymphoma Society translational research grant (NCM).

## Author contributions

JP: envisioned, study design and supervision, method development, experimentation including qPCR, data analysis and interpretation, manuscript preparation; YR: clinical data collection, data curation, performing experiments, data analysis; MS: blood collection, DNA isolation; TB: assisted in sample collection, DNA isolation, data entry; SS, RG: cytological slide review; VM, PPR, AC, SSM, HB: patient selection; MAS provided expert advice, assisted in data interpretation and critical review of the manuscript; PKP, VC: provided resources, multidisciplinary coordination, intellectual discussion, PKP, VC, MAS, YR, PKP, VM,: final editing of the manuscript.

## Conflict of interest

The authors declare no conflict of interest.

## Notes

### Competing Interest Statement

The authors have declared no competing interest.

### Clinical Trial

the study was not clinical trial. So the study was not registered.

### Funding Statement

Department of Health Research,MOHFW, Government of India

### Author Declarations

Study was carried out at Multidisciplinary Research Unit (MRU), Pt. Jawaharlal Nehru Memorial Medical College, Raipur, Chhattisgarh, India, following approval of the protocol by institutional ethical committee, Pt. J.N.M. Medical College, Raipur, India. All human samples were collected following informed consent from the study subjects.

