## Supplementary Documents for "A standalone approach to utilize telomere length measurement as a surveillance tool in oral leukoplakia"

^1^ Multi-Disciplinary Research Unit (MRU), Pt. J.N.M. Medical College, Raipur, Chhattisgarh, India, ^2^Deptt of Pathology, Govt. Medical College, Rajnandgaon, Chhattisgarh, India, ^3^Deptt of Pathology, Pt. J.N.M. Medical College, Raipur, Chhattisgarh, ^4^Deptt of ENT, Dr.B.R.A.M. Hospital & Pt. J.N.M. Medical College, Raipur, Chhattisgarh, India, ^5^Department of Biochemistry, Pt. J.N.M. Medical College, Raipur, Chhattisgarh, India, ^6^Govt. Dental College, Raipur, Chhattisgarh, India, ^7^Regional Cancer Centre, Dr. B.R.A.M. Hospital, Raipur, Chhattisgarh, India, ^8^Harvard (Dana Farber) Cancer Institute, Boston, USA, ^9^Deptt of Obstetrics & Gynecology, Pt. J.N.M. Medical College, Raipur, Chhattisgarh, India, ^10^VA Boston Healthcare System, Boston, USA.

**Short title:** Telomere length in oral leukoplakia

**Correspondence:**

Jagannath Pal

Multi-Disciplinary Research Unit (MRU),

Pt. J.N.M. Medical College,

Raipur, Chhattisgarh, 492001

INDIA

**Supplementary Materials and Methods**

**Relative Telomere length assay by qPCR**

Primer sequences for TL repeat amplification were TelF5-CGGTTTGTTTGGGTTTGGGTTTGGGTTTGGG TTTGGGTT-3, TelR 5-GGCTTGCCTTACCCTTACCCTTACCC TTACCCTTACCCT-3 and primer sequences for single copy gene 36B4 amplification were 36B4F 5-CAGCAAGTGGGAAGGTGTAATCC-3 and 36B4R 5-CCCATTCTATCATCAACGGGTACAA-3(26). Primers were purchased from (Fugene, Bangalore, India). Genomic DNA of all the samples were normalized to 1ng /µl and variation < ± 0.05ng/µl DNA was tolerated. Each qPCR reaction was performed in 20µl volume containing 4ng genomic DNA and 1X Power SYBER green master mix (Thermo scientific). For telomere amplification, reaction mixture contained 1.5µl of each primer(10pm/µl) and 0.4 µl DTT(0.1M) (sigma). For 36B4 amplification, reaction mixture contained 1µl of each primer(10pm/µl). All the PCR reactions (telomere and 36B4) with paired samples and reference DNA were set on a same 96 well qPCR plate ( thermo scientific) in triplicate. A symmetry was maintained on the PCR plate in positioning of the telomere and 36B4 reaction for each sample so that any possible positional effect of the wells could be nullified. PCR was run on a Light Cycler 96 instrument (Roche Life Science). PCR programme was as follows (for both telomere and 36B4 amplicons): 10 min at 95°C, followed by 40 cycles of 95°C for 15 sec, 60°C for 1 min. Standard curve was prepared using serial dilution of Synthetic template for telomeric repeat amplicon (TlS: (TTAGGG)14) and the single copy gene amplicon oligomer(36B4S:5’-CAGCAAGTGGGAAGGTGTAATCC GTCTCCACAGACAAGGCCAGGACTCGTTTG TACCCGTTGATGATAGAATGGG) as described by O’Callaghan. PCR products were checked by running 2% agarose gel electrophoresis and the gel was viewed in chemidoc system ( Syngene ).

**qPCR Data analysis**

Primers efficiency for telo and 36B4 primer pairs were 104.17% and 108.45% respectively. Ct values of the samples were accepted if it were within the linear range of the respective standard curve. Coefficient of variance (CV) within a experiment irrespective of well position in a 96 well PCR plate (margin or internal excluding 4 corner) for Ct values of telomere repeat and 36B4 amplicon were 1.4% and 0.05% respectively. ΔCt was calculated by subtracting the average 36B4 Ct value from the average telomere Ct value using the formula ΔCt = Ct (telomere) – Ct (36B4). The T/S ratio for each sample and reference DNA were calculated using the formula 2^-ΔCt^. For measuring relative TL of buccal epithelim T/S ratio of buccal cells (O) were divided by T/S ratio of paired PBMC (M) or reference DNA (HCT116) (R) which is used as positive control for short TL. The O/M ratio also is referred as ‘TL ratio’ in this manuscript. CV of rTL ratio between the plates was determined by running HCT116 cell line DNA and one of the two batches of cocktail of 5 brush biopsy samples from healthy control as pair in 4 different qPCR plate in different days for each cocktail. CV was 4.5% and 8% for each cocktail.

**Supplementary table 1**

**Test of normality of the samples (Using The Kolmogorov-Smirnov Test of Normality)**

|  | **HC** | | | **OL** | | | | | |
| --- | --- | --- | --- | --- | --- | --- | --- | --- | --- |
|  | **Total(44)** | **NOHC(25)** | **OHC (19)** | **Total (50)** | | **High risk oral habit(47)** | | **No high risk habit (3)** | |
|  |  |  |  | OLN | OLP | OLN | OLP | OLN | OLP |
| K-S test statistic (D) | 0.096 | 0.141 | 0.221 | 0.090 | 0.091 | 0.110 | 0.095 | NA | NA |
| P | 0.777 | 0.653 | 0.27 | 0.780 | .766 | 0.580 | 0.757 | NA | NA |
| Normality | Yes | Yes | Yes | Yes | Yes | Yes | Yes |  |  |

**Supplementary table** 2

**Correlation statistics of TL ratio of Patches (P) and paired distal normal site (N) in OL patients**

| **OL patients** | **Correlation (R)** | | |
| --- | --- | --- | --- |
|  | **Age vs OLN**  **R(p)** | **Age vs OLP**  **R(p)** | **OLN vs OLP**  **R(p)** |
| **Total (50)** | 0.160 (0.268) | 0.090 (0.539) | 0.389 (0.005*) |
| **Habit (47)** | 0.141  (0.343) | 0.037  (0.804) | 0.381  (0.008*) |
| **No hobbit (3)** | NA | NA | NA |

R: The *r* statistics (measure of correlation); *p value is significant at the significance level	p=0.05.

**Supplementary table** 3

**Correlation of TL with duration high risk habit**

| **samples** | **Duration of habit (Y)**  **Mean ±SD (Range)** | **Correlation**  **Duration of habit vs TL : R(p)** | **Correlation**  **Duration of habit vs age : R(p)** |
| --- | --- | --- | --- |
| OL (oral Habit) | 15.47±11.40  (2-50) | OLN: 0.114 (0.449)  OLP: -0.0728 (0.634) | 0.6217 (< .00001) |
| HC (oral Habit) | 10.13 ± 8.343  (2-30) | 0.0177 (0.943) | 0.583 ( 0.0877 ) |

OL: Oral Leukoplakia patients, HC: Healthy control, R: The *r* statistics (measure of correlation); *p value is significant at the significance level	p=0.05.
